# Genome sequencing as a generic diagnostic strategy for rare disease

**DOI:** 10.1101/2023.09.28.23296271

**Authors:** Gaby Schobers, Ronny Derks, Amber den Ouden, Hilde Swinkels, Jeroen van Reeuwijk, Ermanno Bosgoed, Dorien Lugtenberg, Su Ming Sun, Jordi Corominas Galbany, Marjan Weiss, Marinus J. Blok, Richelle A.C.M. Olde Keizer, Tom Hofste, Debby Hellebrekers, Nicole de Leeuw, Alexander Stegmann, Erik-Jan Kamsteeg, Aimee D.C. Paulussen, Marjolijn J.L. Ligtenberg, Xiangqun Zheng Bradley, John Peden, Alejandra Gutierrez, Adam Pullen, Tom Payne, Christian Gilissen, Arthur van den Wijngaard, Han G. Brunner, Marcel Nelen, Helger G. Yntema, Lisenka E.L.M. Vissers

**Affiliations:** Department of Human Genetics, Radboudumc, Nijmegen, Netherlands; Research Institute for Medical Innovation, Radboudumc, Nijmegen, Netherlands; Department of Clinical Genetics, Maastricht University Medical Center, Maastricht, Netherlands; Illumina Inc., Cambridge, United Kingdom

**Keywords:** rare disease, genome sequencing, impact modeling, reducing workflow complexity, genetic diagnostic laboratories, germline variant detection

## Abstract

**Background:** To diagnose the full spectrum of hereditary and congenital diseases, genetic laboratories use many different workflows, ranging from karyotyping to exome sequencing. A single generic high-throughput workflow would greatly increase efficiency. We assessed whether genome sequencing (GS) can replace these existing workflows aimed at germline genetic diagnosis for rare disease.

**Methods:** We performed GS (NovaSeq^™^6000; 37x mean coverage) on 1,000 cases with 1,271 known clinically relevant variants, identified across different workflows, representative of our tertiary diagnostic centers. Variants were categorized into small variants (single nucleotide variants and indels <50 bp), large variants (copy number variants and short tandem repeats) and other variants (structural variants and aneuploidies). Variant calling format files were queried per variant, from which workflow-specific true positive rates (TPRs) for detection were determined. A TPR of ≥98% was considered the lower threshold for transition to GS. A GS-first scenario was generated for our laboratory, using diagnostic efficacy and predicted false negative as primary outcome measures. As input, we modeled the diagnostic path for all 24,570 individuals referred in 2022, combining the clinical referral, the transition of the underlying workflow(s) to GS, and the variant type(s) to be detected.

**Results:** Overall, 95% (1,206/1,271) of variants were detected. Detection rates differed per variant category: small variants in 96% (826/860), large variants in 93% (341/366), and other variants in 87% (39/45). TPRs varied between workflows (79-100%), with 7/10 being replaceable by GS. Models for our laboratory indicate that a GS-first strategy would be feasible for 84.9% of clinical referrals (750/883), translating to 71% of all individuals (17,444/24,570) receiving GS as their primary test. An estimated false negative rate of 0.3% could be expected.

**Conclusion:** GS can capture clinically relevant germline variants in a ‘GS-first strategy’ for the majority of clinical indications in a genetics diagnostic lab.

## Background

Although human genetic diseases are rare, they account for an important public health burden(^1, 2^). Diagnostic approaches to detect the underlying genetic causes of these diseases require a broad spectrum of technologies, ranging from traditional approaches such as karyotyping, genomic microarrays, FISH, and Sanger sequencing, to more advanced technologies, such as exome sequencing and transcriptomics. Each of these technologies is dedicated to detecting one or multiple variant types (3–6). In clinical genomics, (*de novo*) single nucleotide and copy number variants (SNV/CNV) are the most found aberrations(^7–9^), but to a lesser extent aneuploidy, expansions of short tandem repeats (STR), and structural variants (SV) also contribute to disease. To molecularly diagnose a rare disease, multiple workflows are often used, as a single disease can often be caused by multiple variant types(^10–14^). Importantly, for diagnostic purposes, every technology needs to prove clinical, as well as analytical, validity(^3, 15^).

Genome sequencing (GS) promises comprehensive variant calling of all variant types from a single experiment, allowing for all types of molecular diagnoses(^16, 17^). This (potentially) not only leads to an increased diagnostic yield but also provides a higher efficiency for genetic diagnostic laboratories that would no longer need to maintain multiple workflows to capture the various variant types. So far, however, widespread implementation of GS is lagging as the increase in diagnostic yield has been limited while incurring higher costs compared to routine workflows(^18–20^).

A less explored scenario for effective implementation of GS as a routine diagnostic test is the impact of GS replacing currently used diagnostic workflows. For instance, in our tertiary referral centers for genetic diagnostic testing at the Radboud University Medical Center (Radboudumc) and Maastricht University Medical Center+ (MUMC+), approximately 25,000 individuals with a rare disease are tested annually, requiring >10 molecular and cytogenetic workflows to capture all genetic variant types. Replacing these workflows with a single GS-based workflow would increase efficiency. To determine the feasibility of transitioning to a generic GS diagnostic workflow, we performed GS on 1,000 individuals previously molecularly diagnosed with a rare genetic disease, representative of the myriad of genetic variant types identified across 10 different workflows and modeled the impact of a GS-first diagnostic strategy for rare disease in our centers.

## Methods

### Cohort selection

We retrospectively selected archival residual DNA material from a cohort (n=1,000) with known clinically relevant variants (n=1,271) from genome diagnostic laboratories of the Radboudumc in Nijmegen and the MUMC+ in Maastricht. The cohort was representative of the various molecular and cytogenetic workflows used in these departments for the primary diagnosis of germline variants underlying hereditary and congenital diseases using blood-derived DNA (n=979 cases, 1,249 variants; **Supplementary Table S1 and S2**). The cohort was complemented with a few interesting cases for which DNA was extracted from another source than EDTA blood (n=21, 22 variants; **Supplementary Table S1 and S2**). Of note, the cohort included 62 cases with diagnostic referrals that are under suspicion of harboring variants that are at risk to fail detection in a 30x short-read genome. These cases had variants (n=123) in genes or regions with a high level of sequence homology (n=63), or possible mosaic variants (n=60, range 2.4-50%), where the primary diagnostic referral was not always aimed at germline testing, but EDTA blood samples were available (**Supplementary Table S2**).

This study was performed as part of a local validation study for the implementation of GS under ISO15189 accreditation and assessed as a diagnostic innovation by the Medical Ethics Review Committee Arnhem-Nijmegen under dossier number 2020-7142.

### Genome sequencing

GS was performed as defined by the manufacturer (Illumina, San Diego, CA, USA). In brief, 1,000 ng DNA was used for library preparation using the Illumina DNA PCR-free protocol and DNA was tagmented to an average insert size of 450 bp using bead-linked transposomes(^21^). To allow equimolar pooling of samples, barcoded dual indexing was used after which the Illumina index correction strategy was applied (**Supplementary Table S1**). Sequencing was performed on an Illumina NovaSeq6000^TM^ Instrument (24 samples on a S4 flowcell) to an anticipated genome-wide coverage of 30-fold minimal.

### Data analysis

Raw output was stored in Illumina’s BaseSpace Sequence Hub and data was analysed using the Germline Pipeline of Illumina’s DRAGEN^TM^ (Dynamic Read Analysis for GENomics) Bio-IT platform v3.7.5(^22, 23^). In short, after data is demultiplexed, mapped and aligned (GRCh37), the DRAGEN Germline Pipeline provides a comprehensive analysis, including SNV, CNV, SV, as well as repeat expansion detection and genotyping through Illumina Expansion Hunter(24). In addition, we used newly developed DRAGEN SMA(25) and CYP21A2 (DRAGEN v3.9) callers for those specific cases in which the genetic variants located in *SMN1/2* or *CYP21A2* (n=19 cases, 34 variants).

### Variant detection strategy

Variant detection was divided into two phases. First, variant call format files (VCF) generated by the DRAGEN Germline Pipeline were assessed by automated or manual targeted queries using Illumina’s TruSight Software Suite v2.5 (TSS) to identify the variants of interest, resulting in a positive “+” (detected) or negative “-” (not detected) result. Variant detection is based on matching of chromosomal coordinates and alleles for small variants, or reciprocal overlap of genomic event intervals for large variants. Second, variants that failed detection were further assessed to determine why they were absent from the VCFs.

### Sensitivity analysis

Sensitivity analysis was performed in two ways. First, we assessed the overall sensitivity of GS by calculating the true positive rate (TPR) for each workflow, defined as the number of true positive variants (TP) divided by the total number of variants (n=1,271 in 1000 cases) including the false negatives (FN; TPR=TP/(TP + FN). Second, we repeated the analysis after exclusion of the cases (n=62) with variants (n=123) which were *a priori* known to fail detection in a 30x short-read genome to better approximate the TPR.

### Impact analysis

We modeled a scenario of the overall impact of GS implementation as a generic workflow. Hereto we performed three *in silico* analyses.

First, we determined the sequence depth at genomic positions that are known to harbor (likely) pathogenic variation. Sequence depth was calculated from 35 randomly selected genomes. The median coverages were subsequently intersected with genomic positions (coordinates) of all known pathogenic variants reported in the repository of the Dutch Association of Clinical Laboratory Geneticists(^26, 27^) and ClinVar (^28^). In addition, we determined the median coverages for all coding positions of genes with well-established rare disease associations(^29^). Under the assumption that sequence coverage is one of the main determinants for being able to reliably call a variant, we next calculated the fraction of variants with sufficient coverage. Minimal threshold for presumed detection of a variant was set at 10-fold coverage at the respective genomic coordinate. Assuming a binomial distribution with probability 0.5 of sequencing the variant allele at a heterozygous position, at least 10 reads are required to obtain a 99% probability that at least two reads contain the variant allele(^30^).

Secondly, we extrapolated and modeled the obtained workflow based TPRs and GS variant detection limitations from our experimental data to a real-life scenario of our genetic diagnostic laboratories. In line with guidelines for assuring the quality of diagnostic next-generation sequencing (^31, 32^), we used a TPR of ≥98% as threshold for replacing workflows by GS. As input for our model, we used anonymized data of all 41,691 individuals tested in our genetic diagnostic laboratories in 2022 (**Supplementary Figure S1**). For each diagnostic referral (n=54,680; **Supplementary Figure S1**), we evaluated the reason for referral and eligibility for inclusion in our model. A total of 24,166 referrals were excluded, as these either represented cascade screening (n=7,854) or were not within the current scope of replacing by GS (n=16,312), such as for instance non-DNA based and/or biochemical assays (**Supplementary Figure S1**). For the remaining 30,514 referrals for testing, performed in 24,570 individuals, we determined the experiments and workflows used to address the diagnostic referral as input for the model. Combining the workflow and variant type detected per clinical indication, we modelled the impact of substituting eligible experiments (clinical indications) for GS in the diagnostic trajectory of these individuals. Of note, for individuals with multiple referrals that could be replaced by GS, a maximum of one GS was considered, with subsequent diagnostic referrals involving reanalysis of existing data.

Finally, to determine the impact of the GS-first strategy on overall diagnostic yield, the outcome per individual was projected under the following assumptions:

- Negative diagnostic results remained negative, regardless of the underlying workflow, thus also not considering a possible added diagnostic value of GS.
- For individuals whose diagnostic track would not include GS, or where GS was supplemented with an additional non-GS transferable clinical referral, the original diagnostic outcome was maintained.
- For individuals with a conclusive ((highly) likely pathogenic variant), or possible (variant of unknown significance) diagnosis, the GS diagnostic outcome was offset with the TPRs per workflow. Of note, for individuals with multiple diagnostic referrals, it was first determined which experimental workflow led to the initial possible/conclusive diagnosis.

We subsequently determined the number of individuals negatively impacted by the GS-first strategy as proxy for false negatives. The false negative rate (FNR) was determined by FNR=[FN]/[FN]+[TP], in which [TP] was defined as the original diagnostic yield in the cohort of 24,570 individuals minus the [FN].

## Results

### Genome diagnostics and cohort demographics

This local 1000 genome project included archival DNA samples of 505 males and 495 females who were genetically tested in our laboratories using 10 different workflows (**Supplementary Figure S2; Supplementary table S1**). For 378 individuals, this included analysis of specific variants, a single, or a few genes, whereas in 617 individuals, extensive gene panels or other genome-wide analyses were used. For the remaining five individuals, a combination of both approaches was employed (**Supplementary Figure S2**). A total of 1,271 diagnostically relevant variants were reported (**Supplementary Figure S2; Supplementary table S2**). All variants were called complying to specifications of DRAGEN variant calling, grouping them in three categories: a category for small variants (n=860), including SNVs and indels up to 50bp in size, a second one for large variants (n=366), i.e. CNVs and STRs, leaving a third category for all other variants (n=45), involving SVs and chromosome anomalies (CA) (**Supplementary Figure S2; Supplementary table S2**). For our 1000 genomes we reached an average sequencing depth of 37x (**Supplementary Figure S3**).

### GS technical validation and feasibility assessment of replacing workflows by GS

In total, 94.9% (1,206/1,271) of all variants were detected with GS (**Figure 1; Supplementary Table S2**). Small variants were detected in 96.1% (826/860), large variants (123 bp – 72.8Mb) in 93.2% (341/366), and other variants in 86.7% (39/45) (**Supplementary Figure S4**). Subdividing the cohort by the variants we expected to readily identify (n=1,148) and those that we would not (n=123), indeed confirmed the prior knowledge of the technical challenges in detecting mosaic variants and variants located in homologous regions or genes with short-read 30x GS: 1,134 of 1,148 variants (98.9%) were detected as expected, whereas only 72/123 (58.5%) of challenging variants were identified (Fisher’s exact test p<0.001; **Supplementary Table S2**). Of note, the detection limit of small mosaic variants was 13%.

**Figure 1:**
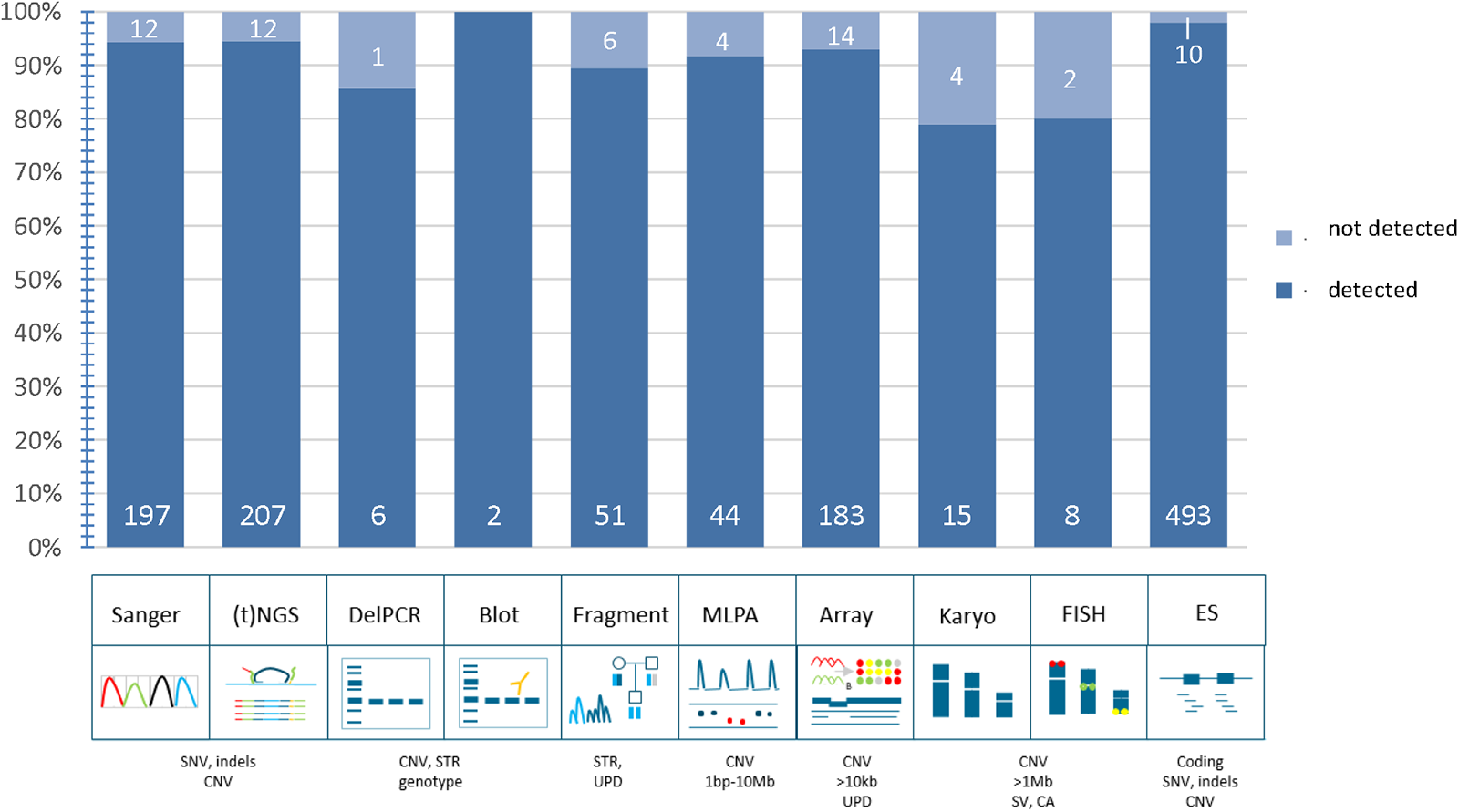
Technical validation of 1,271 variants. Schematic representation of detection rates of previously identified pathogenic variants across multiple different workflows. In total, 94.9% (1,206/1,271) of all variants were detected in GS data. The distribution of variants across the ten workflows shows a detection rate ranging between 79-100%. *Abbreviations: targeted next generation sequencing ((t)NGS), deletion polymerase chain reaction (DelPCR) multiplex ligation-dependant probe amplification (MLPA), fluorescence in situ hybridisation (FISH), exome sequencing (ES), single nucleotide variants (SNV), copy number variants (CNV), short tandem repeat expansions (STRs), uniparental disomy (UPD), structural variants (SV), chromosome anomalies (CA)*

We next reconstituted the 1,271 variants to their original workflows to determine the overall performance of detection of different variant types per workflow, which ranged from 79% for karyotyping to 100% for Southern blots (**Figure 1**). Subsequent analysis of the TPR per workflow revealed that all workflows, except repeat length analysis, karyotyping and FISH, were determined to have a TPR>98% (**Supplementary Table S3**).

### *In silic* of extrapolation of detection rates to 58,393 variants and 4,266 disease genes

Assessing the available coverage data of 794 detected SNVs in our cohort showed that 99.1% had a ≥10x coverage (**Supplementary Table S2; Supplementary Figure S5**). We next leveraged the observations onto a larger in silico data set of variants. Hereto, we obtained 58,393 genomic coordinates from variants known in the VKGL and/or Clinvar databases to cause autosomal dominant/recessive disease (**Supplementary Table S4**) and determined the sequence coverage for those positions across 35 genomes. For 99.5% of variants, the minimal coverage across 35 genomes was ≥10x (**Supplementary Figure S5**). Generation of similar coverage statistics for all coding bases of 4,266 disease-associated genes showed that the average coverage was 45x (**Supplementary Table S5; Supplementary Figure S5**), with 88.1% of genes (3,759/4,266) having a coverage of ≥10x for all protein-coding bases (**Supplementary Figure S5**).

### Modeling the impact of GS implementation in clinical practice

We next set out to model the impact of GS implementation on everyday practice in our clinical centers, from both the clinical point of view, as well as from the laboratory point of view. In addition, we determined the impact on overall diagnostic yield obtained from a GS-first perspective.

In 2022, our tertiary genetic diagnostic laboratory received 30,514 diagnostic referrals to identify the primary germline DNA defect in 24,570 individuals with rare disease (**Figure 2; Supplementary Figure S6**). In total, 883 different reasons for referral were observed, with the top 10 ranking clinical indications being responsible for 21% of all referrals. On average, per individual 1.24 referrals were noted, and 82% of individuals were referred only once (Supplementary Figure S6). Of note, for 966 individuals, the diagnostic referral (n=2,072) consisted of reanalysis of existing exome data and did not require the generation of novel experimental data. For the other 28,442 referrals, 36,633 wet lab experiments were performed using 11 different workflows (**Figure 2**).

**Figure 2:**
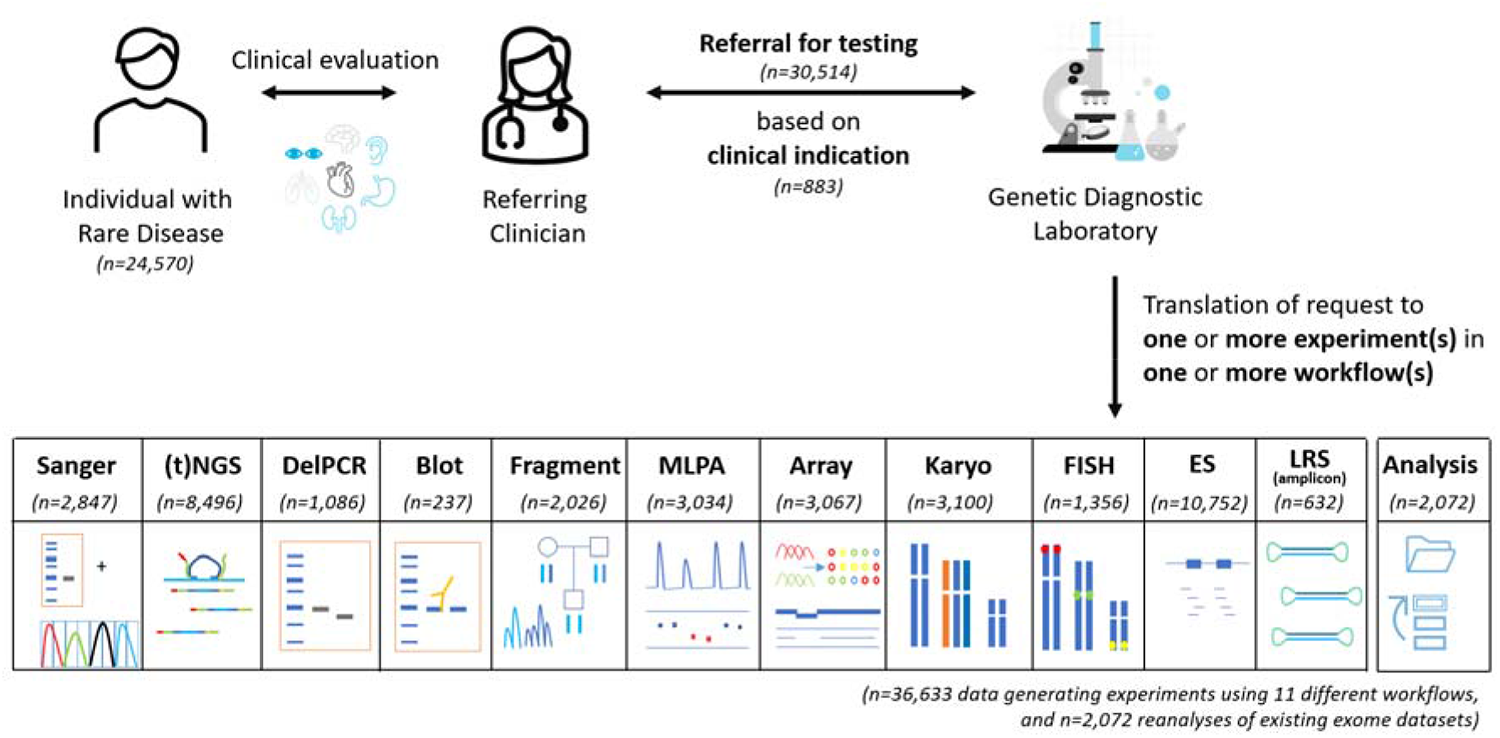
Diagnostic referrals for genetic testing in 2022. In total, 24,570 individuals were referred, together requiring 36,633 data generating experiments (in 23,604 individuals) in 11 different workflows, and 2,072 reanalyses of existing (exome) datasets (in 966 individuals). *Abbreviations: targeted next generation sequencing ((t)NGS), deletion polymerase chain reaction (DelPCR) multiplex ligation-dependant probe amplification (MLPA), fluorescence in situ hybridisation (FISH), exome sequencing (ES), long-read sequencing (LRS)*

From a clinical point of view, 750 of 883 (85%) clinical reasons for referral could be addressed via GS (**Figure 3**). The remaining 133 could not be performed via GS for various reasons, of which somatic variant detection (53%) and detection of variants in homologous regions (13%) are the most prominent (**Figure 3**). From a laboratory point of view, this GS-first strategy would not only fully replace the exome workflow and all Southern blots but would also considerably reduce the use of other workflows, such as Sanger sequencing (by 89%), MLPA (by 80%) and targeted NGS approaches (by 70%; **Figure 3**). Importantly, applying these observations to the diagnostic trajectory of all individuals shows that GS can be used as first-tier test for 16,777 (68%; **Figure 3**) of individuals.

**Figure 3:**
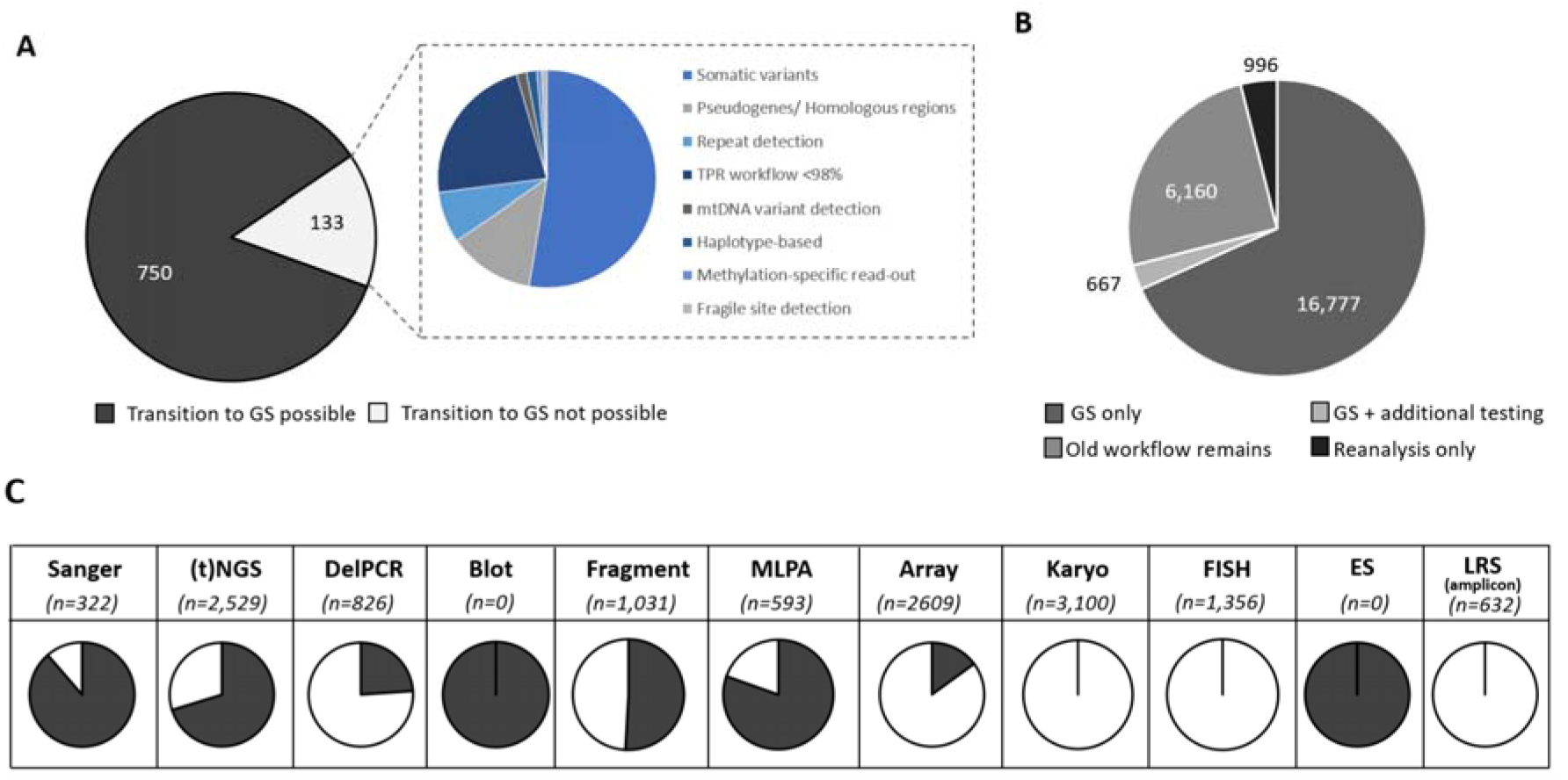
Assessing the impact of a GS-first transition. **A**) From 833 different clinical reasons for referral in 2022, 750 can be transitioned to GS. **B**) This transition would result in 16,777 individuals receiving GS as the only workflow. For 667 (3%), the GS should be supplemented by an additional test, whereas for the remaining 7,126 (29%) GS would not be suited, either because for them the clinical indications included experiments not transferable to GS (n=6,160; 25%), or because the referral did not require data generation (n=966; 4%). **C**) The use of GS as primary test has a significant impact on reducing the experimental workload in the original workflows. Proportions of the transferable number of tests per workflow are indicated in black. *Abbreviations: targeted next generation sequencing ((t)NGS), deletion polymerase chain reaction (DelPCR) multiplex ligation-dependant probe amplification (MLPA), fluorescence in situ hybridisation (FISH), exome sequencing (ES), long-read sequencing (LRS)*

Finally, we modeled the impact on the overall diagnostic yield. In 2022, a conclusive molecular diagnosis was obtained in 2,652 of 24,570 individuals (10.79%), and for another 3,597 (14.64%) a possible diagnosis was identified. Extrapolation of TPRs for individuals whose diagnostic trajectory would include GS, resulted in an anticipated conclusive diagnosis in 2,643 individuals (10.76%) and a possible diagnosis in 3,589 (14.61%; **Supplementary Figure S7**). Collectively, a generic GS-first strategy would thus possibly negatively impact the diagnostic outcome for 17 (0.07%) individuals (FN=17), translating to a possible false negative diagnostic rate of 0.3%.

## Discussion

Over the last decade, the use of GS as a routine diagnostic test has been debated in the context of a higher potential diagnostic yield by interpreting non-coding DNA variants, as well as the potential to diagnose individuals with rare disease more efficiently, as GS allows the identification of virtually all genetic variants in a single experiment. Widespread diagnostic implementation has however been hampered by the costs involved with GS, given that the anticipated higher diagnostic yield has so far not materialized. An increased diagnostic yield is however still expected for unexplained rare genetic disease, especially when looking beyond SNV and CNV detection in the exome only. To ultimately benefit from the advantages of GS, costs need to be reduced for a generic genetic diagnostic laboratory. In this study, we focused on the potential for GS as generic diagnostic rare disease test, replacing the full spectrum of workflows available in a genetic diagnostic laboratory. With our cohort of 1000 genomes, representative of 10 different workflows and a multitude of genetic variant types, we found that GS detected >95% of all pathogenic variants, albeit with variable efficacy across variant types and workflows. We also modeled the impact of a transition to a generic GS workflow for our diagnostic laboratories and conclude that for 68% of individuals diagnostically referred to our departments a generic GS workflow would be possible.

In our series of 1,000 samples, we noted differences in the detection of different variant types; 96.1% of small variants (<50bp) were detected, whereas only 93.3% of large variants, and 86.7% of other variants were recovered from GS. Interestingly, one of the arguments generally used as benefit from GS is its ability to better detect structural variation compared to ES(33). Conceptually, this is true from having a more uniform coverage across the genome(34), and also we, and others, have previously shown that additional diagnoses obtained via GS compared to routine care, are often by structural variants(^35, 36^). However, our data now show that the capture of SNVs/indels from GS is more complete than of structural variants (Fisher’s exact, p=0.006). Another striking observation was the recovery of 72 of 123 variants that we *a priori* expected to be beyond the technical limitations of 30x GS. These included variants located in highly homologous regions such as *STRC* and *OTOA*, as well as variants present in mosaic state (>14%). For the mosaic variants, increasing GS sequence depth may be the only way to recover all clinically relevant variation, especially if present at low variant allele fractions. For capturing variants in homologous regions bioinformatic solutions are under development, allowing the retrieval of (likely) pathogenic variants in these complex genomic regions. Currently, such dedicated callers exist, e.g., we successfully used in our analyses for the SMA(^25^) and *CYP21A2* loci, and for other paralogous regions (37), suggesting that in the near future more (likely) pathogenic variants in such regions can be recovered.

Diagnostic efficacy can be enhanced by reducing the complexity of sample handling and the number of workflows. In our laboratory set-up, one clinical referral is often translated into experiments in multiple workflows; for example, to molecularly diagnose CHARGE syndrome, caused by *CHD7* haploinsufficiency, both Sanger sequencing and MLPA analysis are needed to allow the detection of SNV/indels as well as of (partial) gene deletions. The introduction of a generic GS workflow would allow for calling both SNV/indels, CNVs and other SVs affecting *CHD7* from a single experiment. For other disorders, for instance those caused by the expansion of short tandem repeats, it might be more challenging, as short read sequencing technologies may be unable to capture the full length of the extension. However, our data shows that although for some repeats the exact length cannot be obtained, a generic GS workflow is able to identify those individuals with repeat lengths outside of the normal range. This result can be followed with dedicated tests to determine the size of the repeat. From an efficacy point of view, one may argue that a second workflow is still required. While this is a valid point, in a generic GS workflow, the subsequent use of a second workflow is much more efficient, as it will only be used for those individuals with a high *a priori* chance of a positive outcome (given their abnormal GS results).

Whether or not it is efficient for laboratories to make a transition towards a generic GS workflow may depend on lab-specific factors, including costs, size of the lab, number of workflows in use, and type of diagnostic referrals received. From our series of 1,000 genomes tested, we showed that ES can technically be replaced by GS (TPR>98%), in line with previous reports on comparing diagnostic outcomes of ES and GS (18–20). Hence, diagnostic laboratories, whose expertise is to only perform ES, could easily move towards GS with the benefit of a faster workflow as enrichment is no longer needed(^21^). Yet, for laboratories specialized in the use of karyotyping (TPR<98%) for the detection of somatic copy number changes, routine 30x GS might not be sufficient. The results of our study should therefore be carefully examined and extrapolated to local infrastructure and clinical expertise. Here, we report on our laboratories, which together maintain >10 workflows, representative for most core technologies used in genetic testing(^16^), and enabling detection of all variant types. The scenario models for our centers showed that 750/883 (85%) diagnostic referrals can be completed using GS, which would result in 68% of all individuals referred to our diagnostic laboratory making use of a single workflow and a single experiment, and 3% needing additional testing, suggesting that for 71% of individuals a GS-first strategy would be beneficial. For the remaining 15% of clinical indications not transferable to GS (responsible for 29% of individuals referred), we noted trends, such that most of these required somatic structural variant detection, currently assayed via karyotyping, FISH and/or arrays, or variants were located in complex regions of the genome, currently assessed by amplicon-based long read sequencing strategies(^38^). Based on the results obtained in this study, we could maintain these workflows to be primarily used for these diagnostic referrals. Alternatively, technological innovations specifically targeting these variant types would constitute a worthwhile investment. For somatic variant detection via karyotyping, FISH and/or arrays, optical genome mapping(^39, 40^) could replace these workflows as a second major generic assay, available in parallel to GS, but used for mutually exclusive clinical referrals. Similarly, a more generic use of long read genomes(^41, 42^) may provide a costs-effective strategy for diagnostic referrals involving variants in complex regions in the genome, or where variant size exceeds those detectable from short reads (such as for repeat expansions).

The implementation of a novel technology requires careful balancing of the pros and cons. For GS, our study has highlighted advantages related to laboratory efficiency, but also showed that not all previously detected (likely) pathogenic germline variants were also identifiable from GS. Hence, if a generic GS workflow were to be used, it is to be expected that some individuals who would receive a conclusive diagnosis with the old diagnostic test strategy –, would no longer do so with the implementation of a generic GS. In our objective quantification of the false negative rate from GS, using all diagnoses obtained by the current diagnostic strategy as the gold standard, we modeled that the transition to a generic GS in our laboratory might result in an additional diagnostic false negative rate of 0.3%. Whereas this is undesirable for the individual patient, previous experience has shown that there may be trade-offs. For instance, with the introduction of genomic microarrays at the expense of karyotyping, no longer detecting apparently balanced chromosomal rearrangements had to be accepted. Further, with the introduction of ES as replacement for Sanger sequencing for genetically and clinically heterogeneous disorders, one lost sensitivity at base pair level while gaining in mutation target size. Both innovations changed diagnostic testing, because despite losing out on a few positive diagnoses, they still improved the overall diagnostic yield(^43, 44^). So far, the overall diagnostic advantage of GS is still limited. Disease-specific evaluations of diagnostic yield of GS have, however, reported on an increase in diagnostic yield, ranging from 1.3% for neurodevelopmental disorders(^20^) to 17% for congenital limb malformations(17). Additionally, it has been reported that cytogenetically found apparently balanced chromosomal rearrangements appear to be genomic imbalances in ∼1/3 of patients with *de novo* translocations and inversions(^45, 46^), and that ∼2/3 of balanced chromosomal abnormalities are involved in pathogenic mechanisms(^47^). With growing experience in detecting and interpreting structural variants in GS data, we also expect to identify more inversions, translocations, and other structural variants as underlying causes of human genetic disease. The use of GS over current workflows would provide an added value for which individuals with rare disease would immediately benefit, thus potentially compensating for the 0.3% diagnostic loss from introducing a generic GS workflow.

We note that 6.8% of our referrals (n=2,072) involved reanalysis of existing exome data. With increasing knowledge on the role of (rare) non-coding variants in relation to disease and improvement in the bioinformatic detection of variants in complex regions of the genome from short reads, the availability of GS provides more flexibility in adapting reanalysis strategies towards these loci and variant types in the near future.

## Conclusions

In summary, our study provides detailed insights into the technical possibilities and limitations of GS and its use as generic diagnostic workflow. We show that >95% of known pathogenic variants, selected across the full spectrum of genetic variation, are readily detectable from GS. Modeling the impact of the transition to a generic GS strategy for our laboratory resulted in a more efficient workflow for 71% of individuals by reducing overall test complexity. A possible false negative rate of 0.3% was observed. It is possible that this potential diagnostic loss will be offset by an increase in diagnostic yield expected from GS over standard care, enabled by an evolving GS workflow, guided by better bioinformatic tools to further improve the detection of a wide variety of genomic variants, and a greater understanding of non-coding and structural variant interpretation. GS thus appears a suitable generic first tier test to diagnose individuals with rare diseases.

## Data Availability

The dataset(s) supporting the conclusions of this article is(are) included within the article (and its additional file(s)).

## Abbreviations

CA: chromosome anomaly
CNV: copy number variants
DNA: deoxyribonucleic acid
ES: exome sequencing
FISH: fluorescence in situ hybridization
FN: false negative
FNR: false negative rate
GS: genome sequencing
MLPA: multiplex ligation-dependent probe amplification
NGS: next generation sequencing
SB: Southern blot
SMA: spinal muscular atrophy
SNV: single nucleotide variant
STR: short tandem repeat
SV: structural variant
TP: true positive
TPR: true positive rate
VCF: variant calling format

## Declarations

### Ethics approval and consent to participate

Ethics approval was granted by the Medical Ethics Review Committee Arnhem-Nijmegen (2020–7142). This approval allows for diagnostic follow-up and innovation.

### Consent for publication

Not applicable

### Competing interests

The authors declare that they have no competing interests, aside from Illumina co-authors (X.B., A.P., T.P.) currently being employees and shareholders of Illumina.

### Funding

The study was in part funded through grants from the Dutch Organisation for Health Research and Development (015.014.066 to LELMV). In addition, the aims of this study contribute to the Solve-RD project (to HGB and LELMV), which has received funding from the European Union’s Horizon 2020 research and innovation program under grant agreement No 779257.

### Authors’ contributions

Conceptualization: HB, HY, LV; Data curation: JCG, JP, AG, APu, TP; Formal Analysis: GS, LV; Investigation: GS, RD, AO, HS, JR, ROK, AG, APu, TP; Resources: MN, EB, DL, SS, MW, MB, TH, DH, NL, AS, EK, APa, ML, XZB, CG, AW; Software: JR, JCG, XZB, JP, AG, APu; Supervision: HB, HY, LV; Visualization: GS, LV; Writing – original draft: GS, LV; Writing – review & editing: CG, HB, HY, LV, GS. All authors have read and approved the final version of the manuscript.

## Acknowledgements

Part of the reagents and software pipelines required for the study were kindly provided to us by Illumina. We are most grateful to our Genome technology center for sequencing the genomes and our bioinformatic team for data processing and storage.

## Notes

### Author Declarations

Medical Ethics Review Committee Arnhem-Nijmegen of Radboudumc gave ethical approval for this work

