## Supplemental material for "Genome sequencing as a generic diagnostic strategy for rare disease"

[**Supplementary Table S3:** GS sensitivity](#_Toc143873984)	9

[**Supplementary Table S4:** Online excel file providing Online excel file providing genomic coordinates [GRCh38] and coverage statistics for 58,393 variants for which previously (likely) pathogenic variants were described.](#_Toc143873983)

**Supplementary Figure S1: Scenario model to determine individuals eligible for GS-first strategy**

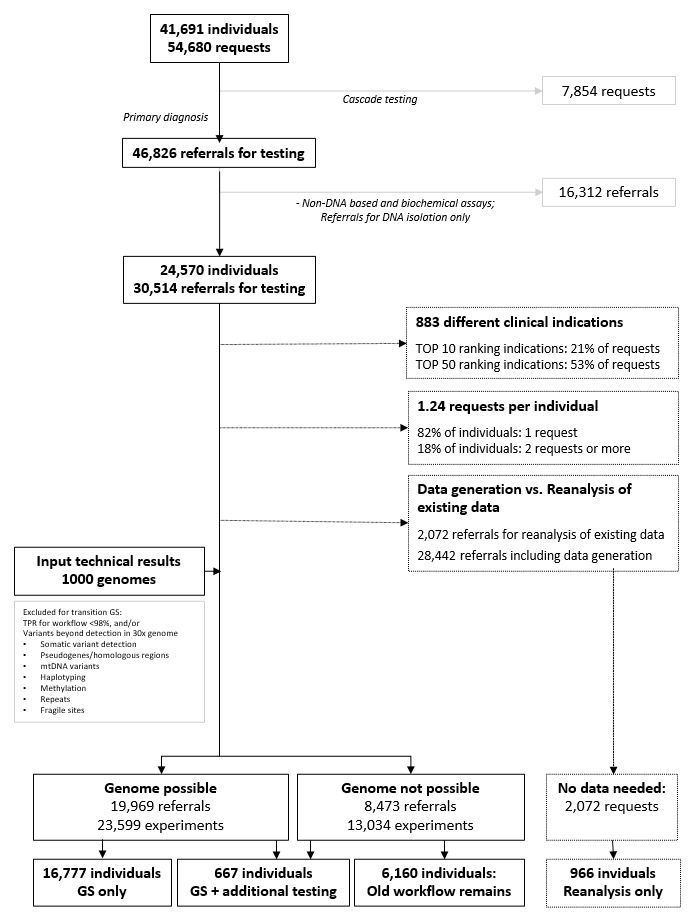

### **Supplementary Figure S2: A cohort of 1000 cases with clinically relevant variants spanning the broad range of genome diagnostics.**

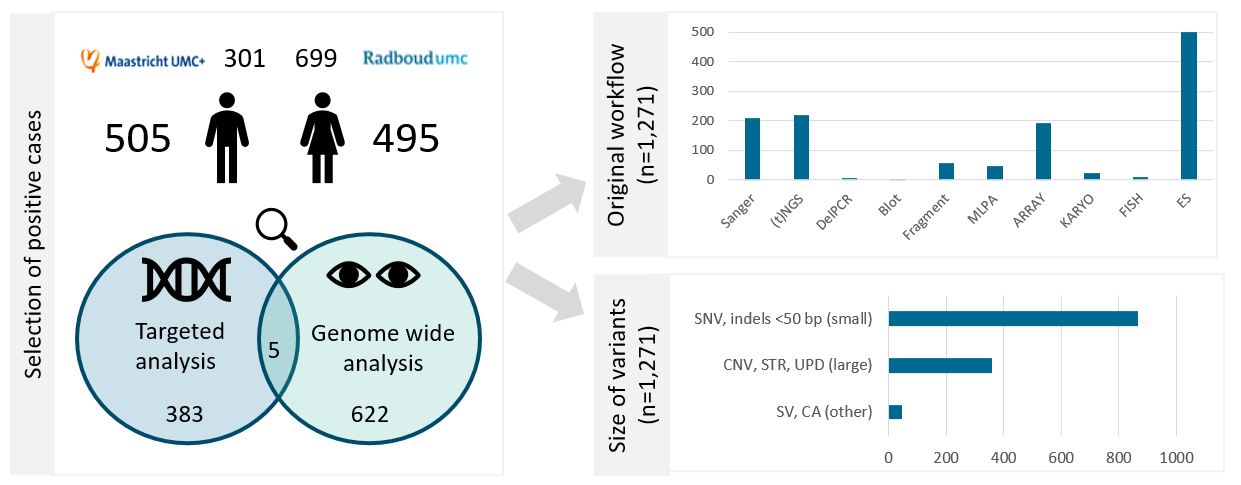

A

B

C

**A** The 1000 genomes cohort consisted of 505 males and 495 females, who were genetically diagnosed in the Radboudumc or Maastricht UMC+ in 2018. The assays that were performed to find these diagnoses were either targeting specific variants and single (or a small set of) genes or complete gene panels or chromosomes were analyzed based on the patient’s phenotype. **B** In these cases, a total of 1,271 variants were identified, requiring >10 different workflows to diagnose them. **C** The variants were grouped in small (<50 bp), large (50 bp and up), and other variants (SVs and CA).

Abbreviations: targeted next generation sequencing ((t)NGS), deletion polymerase chain reaction (DelPCR), multiplex ligation-dependent probe amplification (MLPA), fluorescent in situ hybridisation (FISH), exome sequencing (ES), single nucleotide variants (SNV), copy number variants (CNV), short tandem repeat expansions (STRs), uniparental disomy (UPD), structural variants (SV), chromosome anomalies (CA)

### **Supplementary Figure S3: The average output of 1000 genomes.**

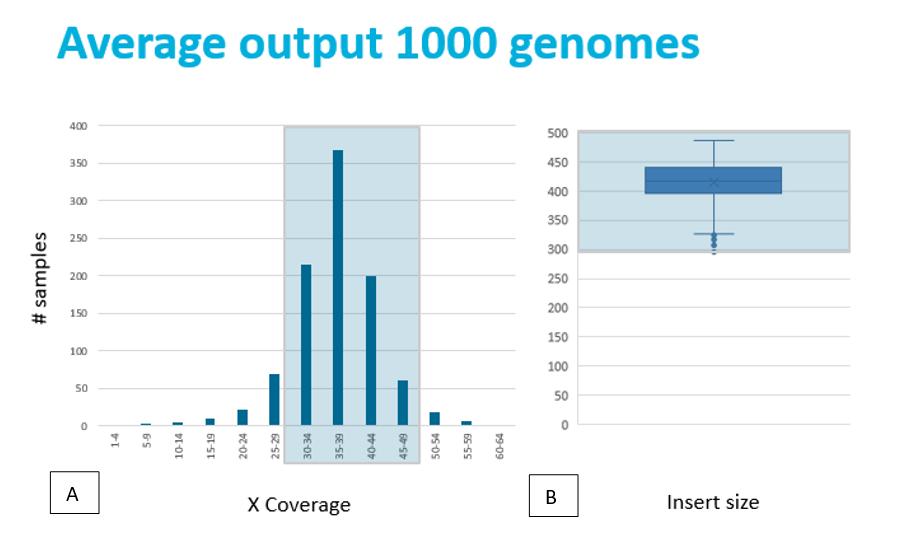

**A** As multiple observations per base are needed to come to a reliable base call, the recommended sequencing depth for genome sequencing is 30x to 50x. **B** Insert sizes are also important for the sequencing. For efficient sequencing, small insert sizes (risk of overlapping paired sequences) as well as larger fragments (decrease of cluster efficiency) must be avoided. We therefore aimed for a 300-500bp range for our 2x150bp paired-end sequencing. In this project we reached an average sequencing depth of 37x and an insert size of around 400-450bp.

### **Supplementary Figure S4: GS Technical validation by variant size and assessment of why variants were not identified**

## **A**

|  | + | - |
| --- | --- | --- |
| CA | 28 | 2 |
| CNV | 262 | 18 |
| ROH, UPD | 26 | 1 |
| SNV, indel | 827 | 34 |
| STR | 52 | 6 |
| SV | 11 | 4 |

**B**

|  | Totaal |
| --- | --- |
| mosaicism | 27 |
| homologues region | 25 |
| repetitive region | 10 |
| other | 3 |

**A** In total, 94.9 % (1,206/1,271) of all variants were detected with GS. Small variants (<50bp) were detected in 96.1% (833/867), large variants (123 bp - 72.8Mb) in 93.3% (334/359), and other variants in 86.7% (39/45). The total list of variants and whether they were present in the GS data (‘detected’ vs. ’not detected‘) can be found in **Supplementary Table S2**. +: detected; -: not detected. **B** In the 5% undetected variants (N=65), we identified common themes that are attributable to short-read 30x GS and downstream analysis. Undetected variants were mostly found in mosaic cases (n=27, 2.4-20%), homologues regions (n=25), i.e. pseudogenes or paralogues genes, or likewise in repetitive regions (n=10), i.e. repeats, telomeres or centromeres.

### **Supplementary Figure S5: *In silico* coverage statistics at variant level and disease genes**

***A:Coverage statistics for 794 detected SNVs from the 1000 Genomes***

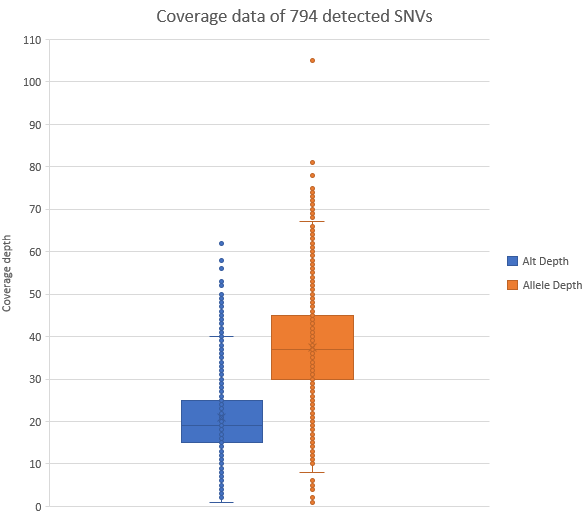

***B: Coverage statistics of 58,393 ClinVar and VKGL variants***

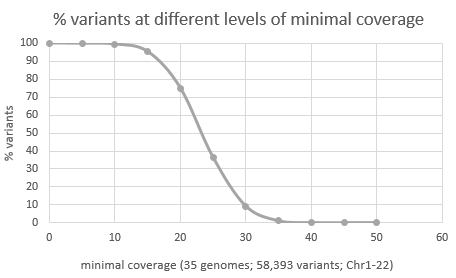

***C-D: Coverage statistics of 4,266 disease genes***

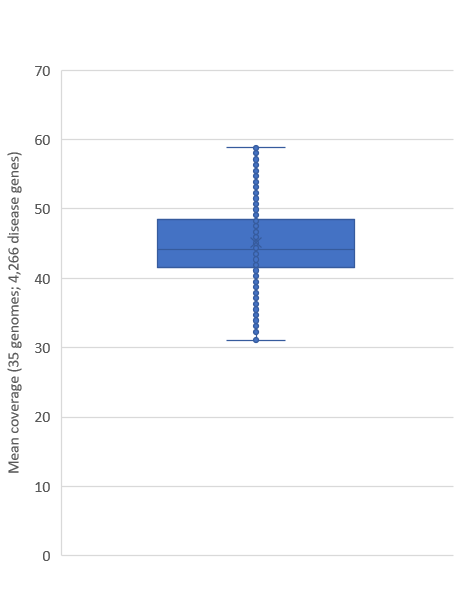

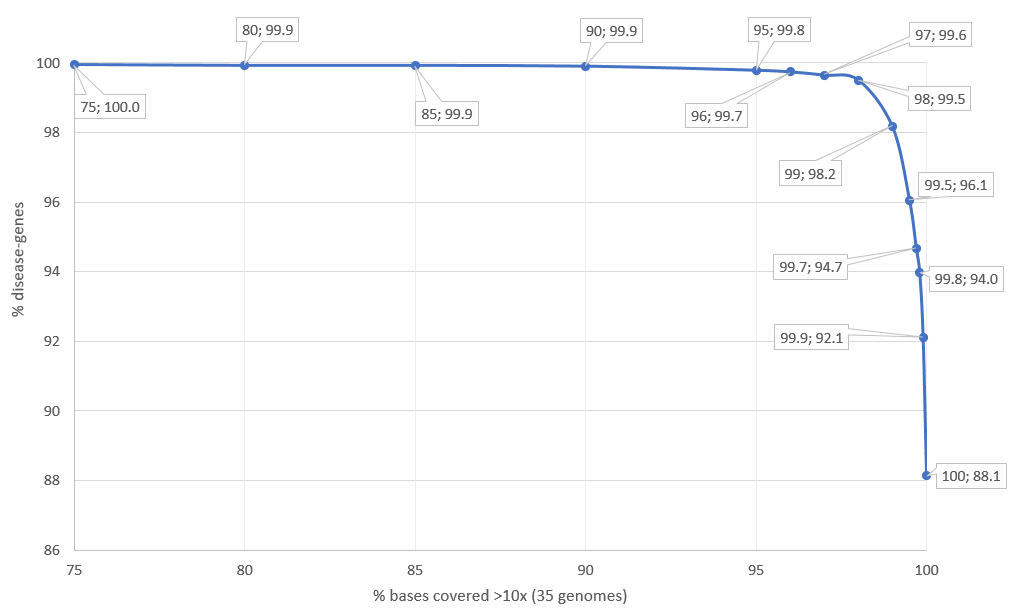

**A** Coverage data of 794 detected SNVs in our cohort, where allele depth ranged from 1-105, and (variant) alternative allele depth ranged from 1-62, with a 13-100% variant range. **B** Sequence depth at genomic positions that are known to harbor (likely) pathogenic variation and **C** Mean coverages for all coding positions of genes with well-established rare disease associations were calculated from 35 randomly selected genomes. **D** The fraction of genes versus the percentage of bases of the gene with ≥10x coverage.

### **Supplementary Figure S6: Schematic representation of referrals to Radboudumc and MUMC+ in 2022**

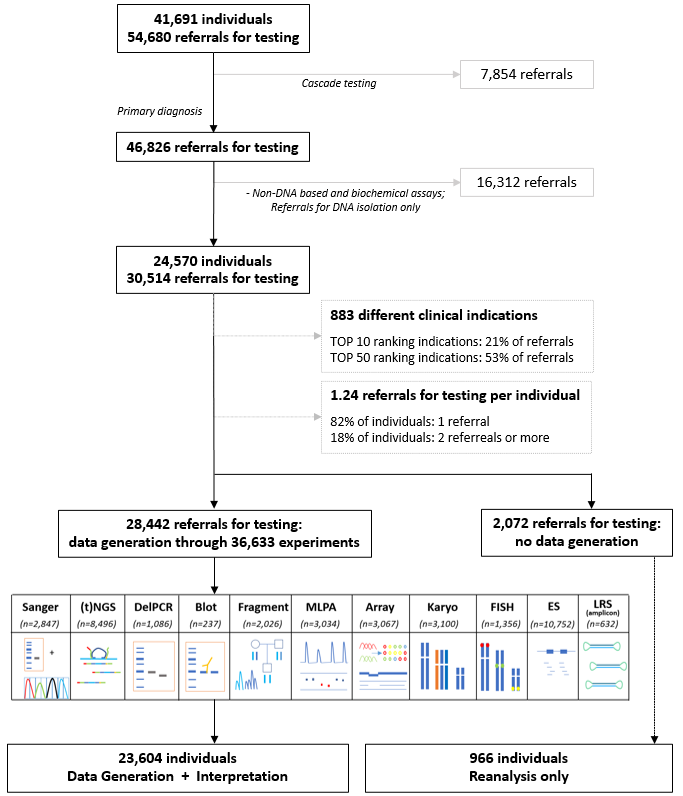

### **Supplementary Figure S7: Schematic overview of assumptions made to evaluate the impact on diagnostic yield from transition to a generic GS approach**

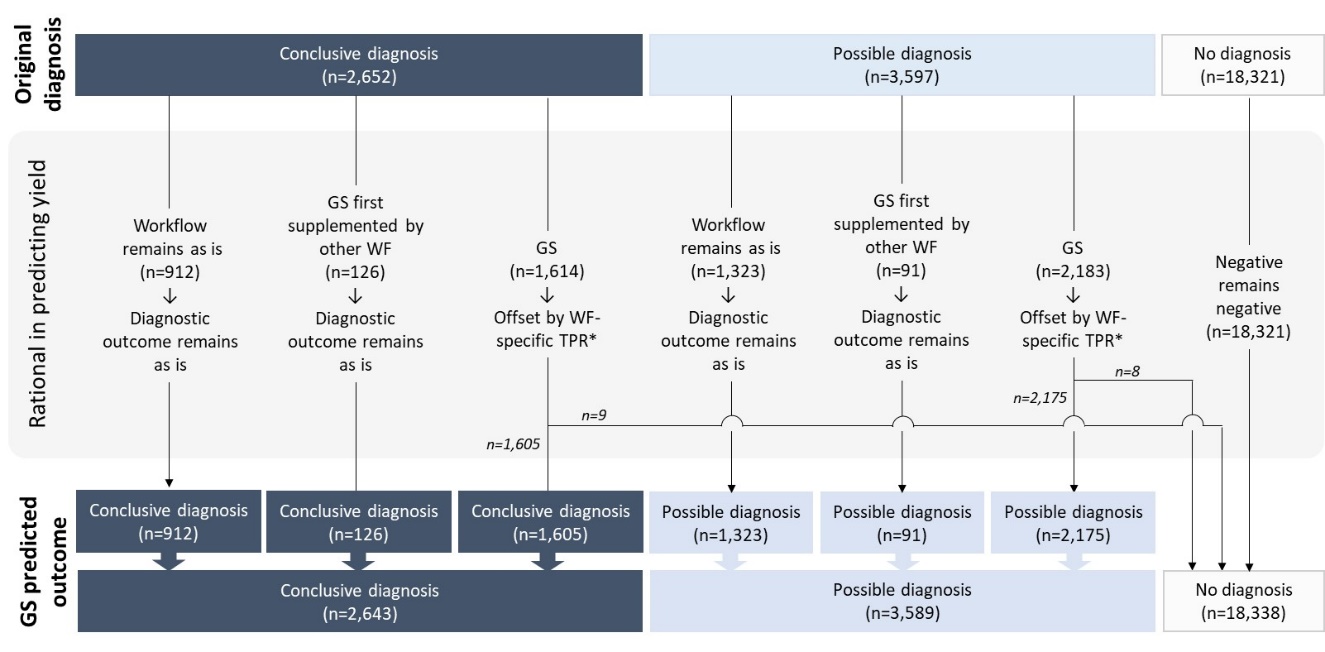
**A**

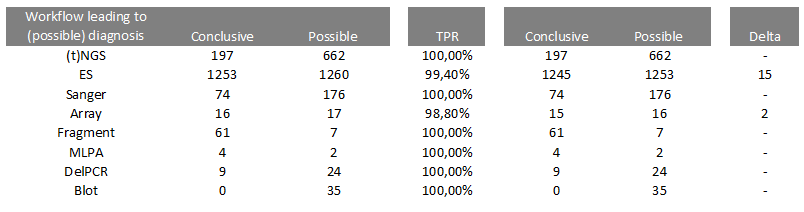

**B**

**A**) Based on clinical referrals being transferable to generic GS, the impact on diagnosis was evaluated for all 24,570. Top row shows original diagnosis per individual, where ‘*n*’ refers to number of individuals; *Offset with workflow specific TPRs are provided in **B**. Assuming all negative diagnoses remain negative, this translates to a possible false negative diagnostic rate of 0.3% (17/6232).

### **Supplementary Table S3: GS sensitivity**

*Excluded indications: Adenomatous polyposis coli, Chronic lymphocytic leukemia, PTEN Hamartoma tumor syndrome (diagnostic referrals that are under suspicion of harboring mosaic variants and/or added to include mosaic variants although not primarily aimed at germline testing); Excluded variants: mosaic variants <20%, variants in the *CYP21A2*, *SMN1*, *OTOA*, *STRC* or OPSIN genes.
TPR≥98% indicated by grey marking

| **# variants** | **1) Technical validation** | | | | **2) Technical validation**  **+ exclusion expected false negatives*** | | | |
| --- | --- | --- | --- | --- | --- | --- | --- | --- |
| **Workflow** | **positive** | **false negative** | **total** | **TPR** | **positive** | **false negative** | **total** | **TPR** |
| Sanger | 197 | 12 | 209 | 94.3% | 178 | 0 | 178 | 100.0% |
| (t)NGS | 207 | 12 | 219 | 94.5% | 191 | 0 | 191 | 100.0% |
| DelPCR | 6 | 1 | 7 | 85.7% | 4 | 0 | 4 | 100.0% |
| Blot | 2 | 0 | 2 | 100.0% | 2 | 0 | 2 | 100.0% |
| Fragment | 51 | 6 | 57 | 89.5% | 51 | 6 | 57 | 89.5% |
| MLPA | 44 | 4 | 48 | 91.7% | 33 | 0 | 33 | 100.0% |
| Array | 183 | 14 | 197 | 92.9% | 168 | 2 | 170 | 98.8% |
| Karyo | 15 | 4 | 19 | 78.9% | 15 | 1 | 16 | 93.8% |
| FISH | 8 | 2 | 10 | 80.0% | 8 | 2 | 10 | 80.0% |
| ES | 493 | 10 | 503 | 98.0% | 488 | 3 | 491 | 99.4% |
| **Total** | **1206** | **65** | **1271** | **94.9%** | **1138** | **14** | **1152** | **98.8%** |
| **Type variant** |  |  |  |  |  |  |  |  |
| SNV, indels | 827 | 34 | 861 | 96.1% | 789 | 3 | 792 | 99.6% |
| STR | 52 | 6 | 58 | 89.7% | 52 | 6 | 58 | 89.7% |
| ROH, UPD | 26 | 1 | 27 | 96.3% | 24 | 0 | 24 | 100.0% |
| CNV | 262 | 18 | 280 | 93.6% | 239 | 2 | 241 | 99.2% |
| CA | 28 | 2 | 30 | 93.3% | 23 | 0 | 23 | 100.0% |
| SV | 11 | 4 | 15 | 73.3% | 11 | 3 | 14 | 78.6% |
| **Total** | **1206** | **65** | **1271** |  | **1138** | **14** | **1152** |  |

*Abbreviations:* *targeted next generation sequencing ((t)NGS), deletion polymerase chain reaction (DelPCR) multiplex ligation-dependent probe amplification (MLPA), fluorescence in situ hybridisation (FISH), exome sequencing (ES), single nucleotide variants (SNV), copy number variants (CNV), short tandem repeat expansions (STRs), uniparental disomy (UPD), structural variants (SV), chromosome anomalies (CA)*
